# ScanTecc classifies primary cancers via cell-free extrachromosomal circular DNA in peripheral blood

**DOI:** 10.1101/2022.06.24.22276859

**Authors:** Jingwen Fang, Yehong Xu, Jing Wang, Benjie Shan, Kaiyang Ding, Shouzhen Li, Qiaoni Yu, Wen Zhang, Yunying Shao, Jiaxuan Yang, Ke Liu, Guangtao Xu, Xinfeng Yao, Ruoming Sun, Mengyuan Zhang, Zhihong Zhang, Yueyin Pan, Chuang Guo, Kun Qu

**Author notes:** Corresponding should be addressed to Kun Qu; Chuang Guo;. These authors contributed equally to this work (Jingwen Fang, Yehong Xu, Jing Wang, Benjie Shan, Kaiyang Ding).

## Abstract

Extrachromosomal circular DNA (eccDNA) is gaining substantial attention in basic and clinical research for its contribution to tumor heterogeneity, biogenesis, and evolution. However, its presence and molecular features in peripheral blood from cancer patients are poorly understood. Here, we demonstrated the existence of eccDNA molecules in cancer patients’ plasma using atomic-force microscopy and high throughput sequencing technology. We then developed a method named ScanTecc (**S**creening **can**cer **T**ypes with cell-free **ecc**DNA) that integrates cell-free circular DNA sequencing and machine learning approaches for cancer patient screening and classification. We applied ScanTecc on a total of 413 patients with multiple types of human primary cancers and 52 healthy individuals and found a significantly greater number and more extended size of eccDNAs in patients’ peripheral blood. ScanTecc accurately distinguished cancer patients (including early stages I and II) from healthy individuals with an overall prediction rate of 0.85 and identified distinct cancer types with an overall prediction rate of 0.84. Our study provides evidence for a non-invasive approach potentially applicable for early detection of cancer patients in clinics.

## Introduction

With the rapid development of sequencing and imaging technology, circular DNA was broadly identified in normal tissues and cancer^1,2^, and thereby is gaining substantial attention from biological and clinical researchers in recent years^3,4^. Circular DNA is commonly recognized in two independent classes: (1) large size and copy number–amplified extrachromosomal circular DNA (referred to ecDNA); (2) small extrachromosomal circular DNA (including microDNA; referred to eccDNA). Recent studies have shown that ecDNA amplification is prevalent in multiple human cancers and is associated with accelerated cancer evolution and poor clinical prognosis^5-7^. This might be attributed to increased expression of oncogenes that are frequently amplified on ecDNAs^8-11^. The circular nature of ecDNA exhibits unique properties, such as high mobility^12^, clustered somatic mutations^13^, and uneven segregation behavior^14^. Thus, ecDNA can form hubs and function as mobile enhancers for oncogenes on other ecDNAs or chromosomes^12,15^. By contrast, eccDNA has been reported as apoptotic products with strong immunostimulatory activity, which is independent of eccDNA sequences but dependent on eccDNA circularity and related cytosolic DNA sensors^16^. Importantly, the amount of both ecDNA and eccDNA in cancer was significantly higher than that in normal tissues^5^, indicating that circular DNA may be a potential biomarker for the diagnosis and treatment for cancer patients.

Liquid biopsy is an emerging technology for cancer diagnosis^17^. Cells from tumor tissues release circulating tumor DNA (ctDNA) due to apoptosis, autophagy, and a number of other mechanisms^18^. The mutation and epigenetic modifications of these ctDNAs provide broad clinical applicable values for cancer screening and treatment^19^. Several studies have reported the presence of cell-free eccDNA in circulation^20-23^, and human eccDNAs were also discovered in the peripheral blood of patient-derived xenograft (PDX) mouse models^19^, prompting eccDNA as a new non-invasive biomarker for diagnosis. Moreover, plasma samples collected prior to surgery are enriched in longer size of eccDNAs compared with the samples from the same patient obtained 6 weeks after surgery, suggesting that longer size of eccDNAs in circulation may be released from tumor cells^20^. Since eccDNA is expected to be more stable compared to linear DNA, it provides an unique advantage for using eccDNA as a diagnostic target for liquid biopsy^24^. However, the current lack of data for characterizing eccDNAs in peripheral blood from cancer patients, especially in a large cohort of cancer patients, has thus limited our capability of applying eccDNA signatures for non-invasive disease diagnosis.

In this study, we investigated the presence and molecule features of eccDNAs in peripheral blood from a large cohort of 413 patients with multiple types of primary cancers and 52 healthy individuals. We demonstrated the presence of eccDNA molecules in both healthy individuals’ and cancer patients’ plasma of peripheral blood by atomic-force microscopy and next-generation sequencing technologies. We further developed a new approach for screening cancer types with cell-free eccDNA (ScanTecc), which integrates cell-free eccDNA profiling, annotation, and biomarker classification using next-generation sequencing and machine learning technologies. Employing ScanTecc in this cohort, we found a greater number and longer size of eccDNAs in plasma from cancer patients than those from healthy individuals. In addition, the size ratio and distribution of these eccDNAs can be used to accurately distinguish cancer patients (including early stages I and II) from healthy individuals with an overall prediction rate of 0.85. We further applied ScanTecc to distinguish major cancer types based on eccDNA gene features and obtained an overall prediction rate of 0.84. Our study provided evidence for the presence of eccDNAs in peripheral blood using a large cohort of patients with multiple primary cancers, leading to the feasibility of applying eccDNA signature for non-invasive cancer diagnosis.

## Results

### Identification of eccDNA in peripheral blood from patients with cancer

To determine whether eccDNAs are present in peripheral blood, we presented ScanTecc, an approach for screening cancer types with cell-free eccDNA. ScanTecc integrates eccDNA profiling with next-generation sequencing, bioinformatics method for eccDNA identification, downstream analyses to identify the junctions created by the circularization of genomic segments, and machine learning algorithms for biomarker identification and cancer screening (Fig. 1A). We first assessed the performance of two widely used but distinct experimental methods (*i*.*e*., whole-genome sequencing data analyzed by AmpliconArchitect algorithm^1^; and Circle-seq^25^) for eccDNA purification and identification. By comparing the efficiency of these two approaches in parallel with the same amount of input cell-free DNA in 3 samples, we found that Circle-seq identified an average of 3521 eccDNAs per sample, while only one circular DNA was identified in 1 out of the 3 samples by WGS combined AA strategy (Supplementary Fig. 1A), which is also confirmed by Circle-seq (Supplementary Fig. 1B). This result revealed the high efficiency and sensitivity of Circle-seq for cell-free eccDNA purification and identification.

**Fig. 1.**
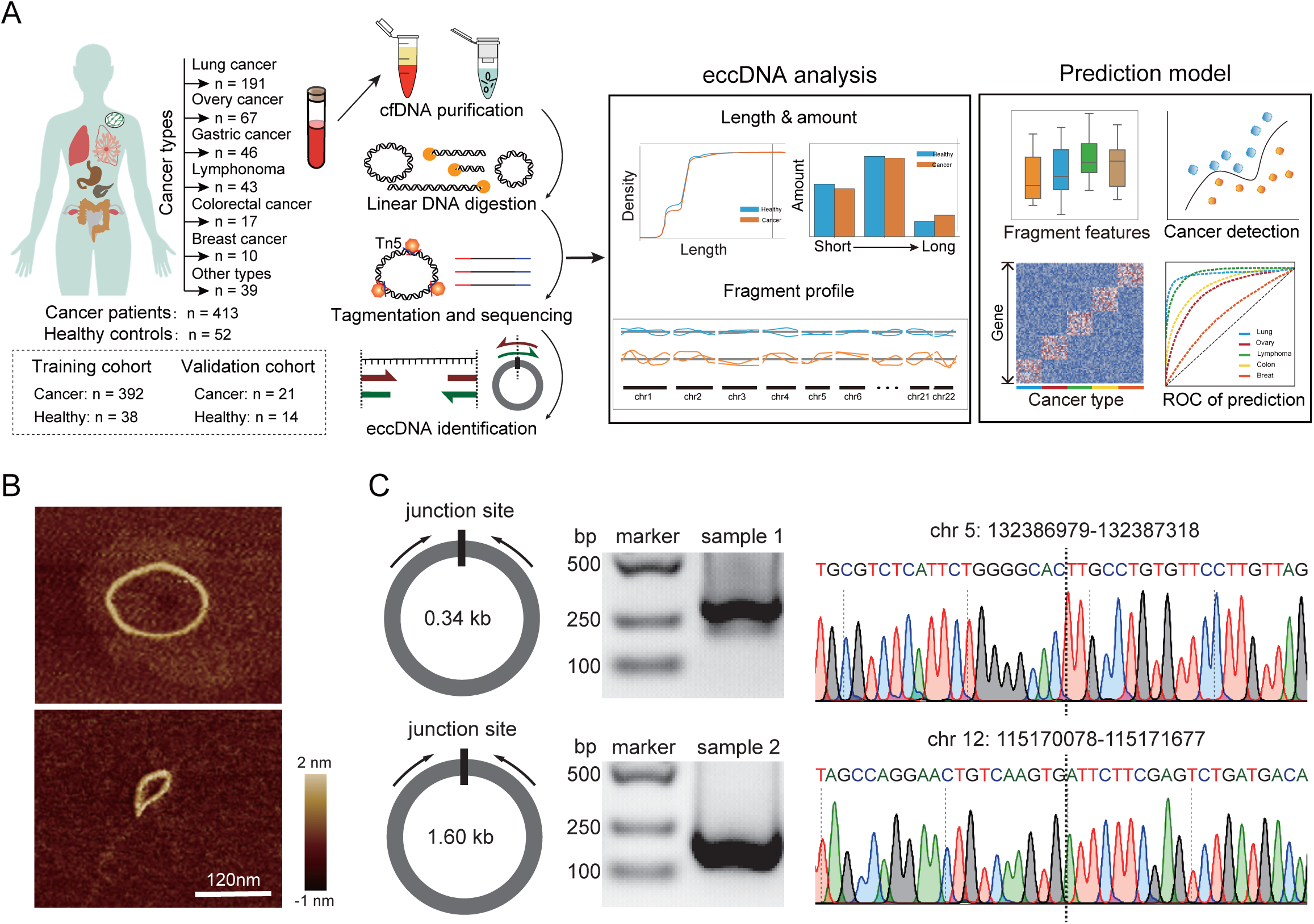
ScanTecc identified cell-free extrachromosome circular DNA. a. Schematic overview of ScanTecc design, including: 1) cell-free eccDNA purification and sequencing; 2) eccDNA identification through split-reads and annotation; 3) machine learning model for distinguish healthy individuals between cancer patients, and identification the tumor of origin b. Cell-free eccDNA purified from blood sample scanned with SAFM. Scale bar, 120 nm. c. PCR & Sanger-sequencing validation of cell-free eccDNA junction site.

To further validate the existence of eccDNAs in peripheral blood, we incorporated an optimized Circle-seq protocol into ScanTecc and purified cell-free eccDNA from 100μg cell-free DNA. We successfully confirmed the purity and circularity of cell-free eccDNAs by scanning atomic-force microscopy (Fig. 1B). We also validated 10 junction sites of cell-free eccDNAs, which were confirmed in plasma from patients with cancer by outward PCR and Sanger sequencing (Fig. 1C). In sum, these results demonstrate that ScanTecc can effectively enrich and identify cell-free eccDNA in peripheral blood.

### ScanTecc identified eccDNAs from peripheral blood in a large cohort of patients with multiple cancer types

To explore the landscape of eccDNAs from peripheral blood in multiple human cancer types, we obtained 465 high-resolution eccDNA profiles from peripheral blood samples in patients with primary cancer and healthy individuals using ScanTecc. Specifically, our dataset was generated from 413 patients with primary cancer, including lung (n = 191), ovarian (n = 67), gastric (n = 46), lymphoma (n = 43), colorectal (n = 17), breast (n = 10), and the other type of primary cancers (n = 39), as well as 52 healthy individuals (Fig. 1A). Among them, 392 cancer patients and 38 healthy individuals were set as the training cohort and 14 healthy samples and 21 lung cancer samples were collected as a validation cohort (Fig. 1A). None of the patients with cancer had undergone previous treatment (Table S1).

Each eccDNA library was sequenced to yield an average of approximately 83 million reads that are mapped to the human genome (Supplementary Table S1). With this dataset, we found that each blood sample could detect an average of 8949 eccDNAs. Consistent with previous studies on eccDNA profiles from maternal plasma and urine^22,23^, the majority of eccDNAs (∼80%) from peripheral blood samples were smaller than 500bp, suggesting that most of the circular DNA in peripheral blood are microDNAs. The identified eccDNAs were mostly at ∼200 and ∼350 bp in terms of their length (Supplementary Fig. 2A). We further identified that the absolute number of eccDNAs from cancer patients was significantly higher than that from healthy individuals (Fig. 2A and Supplementary Fig. 2B). In particular, when we calculated the number of eccDNA fragments in each 10-fold length bin from 10^0^ bp to 10^8^ bp and constructed an eccDNA length distribution profile for each sample, we found that cancer patients were enriched with longer eccDNAs (>1000bp) in their peripheral blood, while the healthy individuals were enriched with shorter eccDNA (Fig. 2B).

**Fig. 2.**
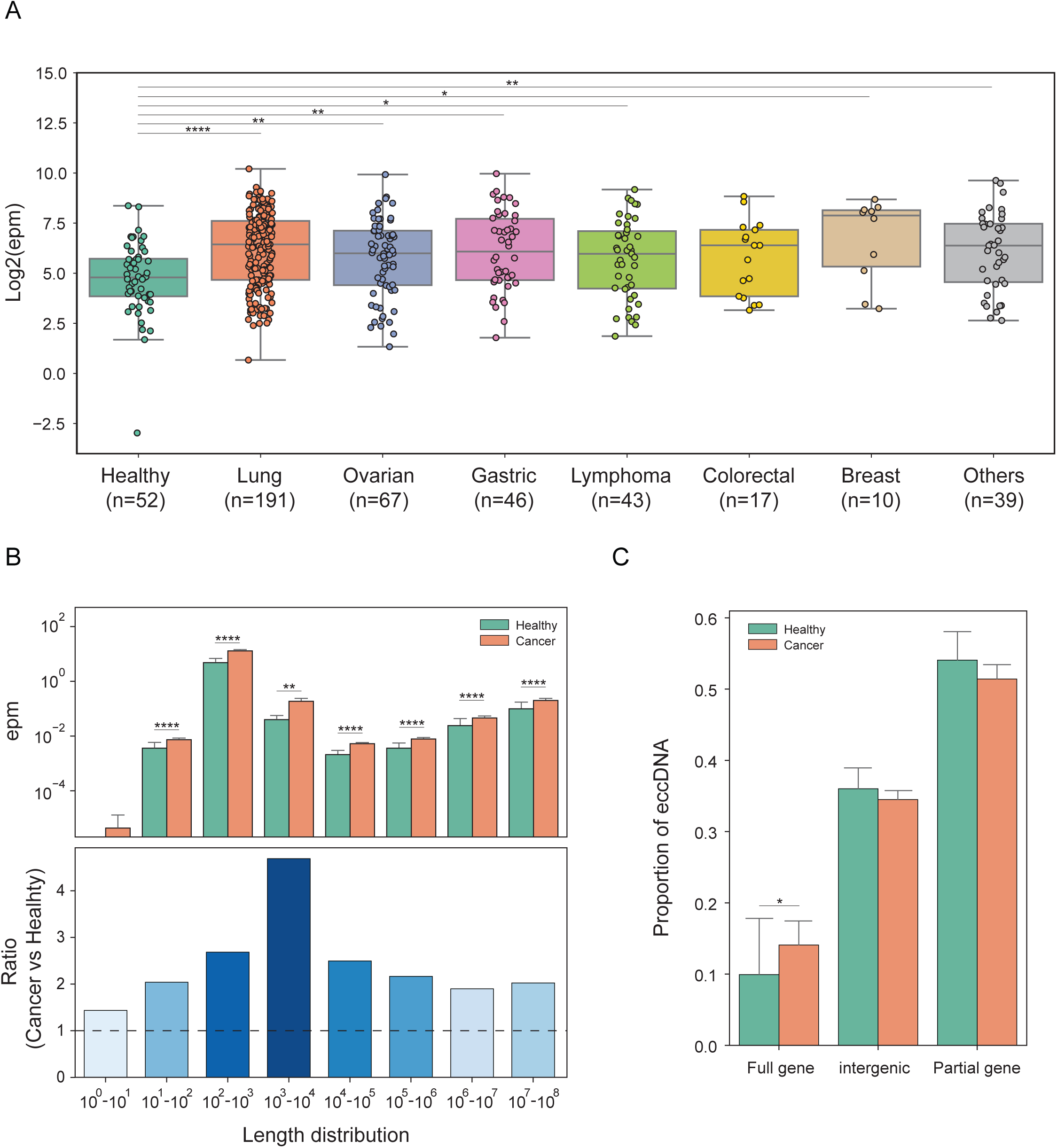
Differential cell-free eccDNA profiles in cancer patients and healthy individuals. a. Normalized cell-free eccDNA counts detected in healthy individuals’ group and 8 cancer types patients’ group. b. Normalized cell-free eccDNA counts detected in healthy individuals’ group and cancer patients’ group 10-fold length bin (upper). Bar plot of Log2 fold-change between patients and healthy individuals in each bin, normalized by log2 fold-change of total counts (lower). c. Fraction of genomic regions covered by patients’ cell-free eccDNA compared to healthy individuals’ cell-free eccDNA.

We then investigated the molecular features of the eccDNAs identified from cancer patients’ plasma and found that they were enriched in the intergenic regions, with relatively lower frequency in the 5’-untranslated regions (5’-UTRs) (Supplementary Fig. 2C). Compared with those from the healthy individuals, eccDNA molecules in peripheral blood from cancer patients were enriched with full-length genes (Fig. 2C), which was consistent with the fragment distribution of the observed circular DNA in tissues from previous studies^26^, suggesting that cell-free eccDNA identified in peripheral blood may be released from tumor tissues. However, there is no significant difference between cancer types (Supplementary Fig. 3A). In addition, our results showed that both the start and end positions of eccDNA molecules were flanked by 5’ A/3’ T nucleotide; besides, no significant difference was observed in nucleotide motif enrichment of eccDNAs between cancer patients and healthy individuals (Supplementary Fig. 2D).

### ScanTecc identifies cancer patients from healthy individuals through eccDNA size and fragment distribution

Given that genome-wide analyses of cfDNA fragmentation were state-of-the-art approaches for non-invasive cancer detection^27,28^, we sought to investigate cell-free eccDNA fragment distribution in cancer patients and healthy individuals (Supplementary Fig. 4A). We first divided the genome into 10Mbp windows. Within each window, we defined the logarithm of the ratio of the numbers of the long (280-500bp) to short (150-280bp) fragments as the fragment profile, and calculated the fragment profiles for each of the 413 cancer patients and 52 healthy individuals (Fig. 3A). An analysis of the commonly altered genomic windows highlights the position-dependent alterations in eccDNA fragments, revealing 14%-38% affected windows across the cancer types (Fig. 3B). We then performed pair-wise Pearson-correlation analyses of the fragment profiles for each sample, and we found that similar to cfDNA fragmentation distribution in patients with cancer^29^, the median correlation scores of cancer patients were significantly lower than those of the healthy individuals (Fig. 3C, *P* < 0.0001, Student’s t-test).

**Fig. 3.**
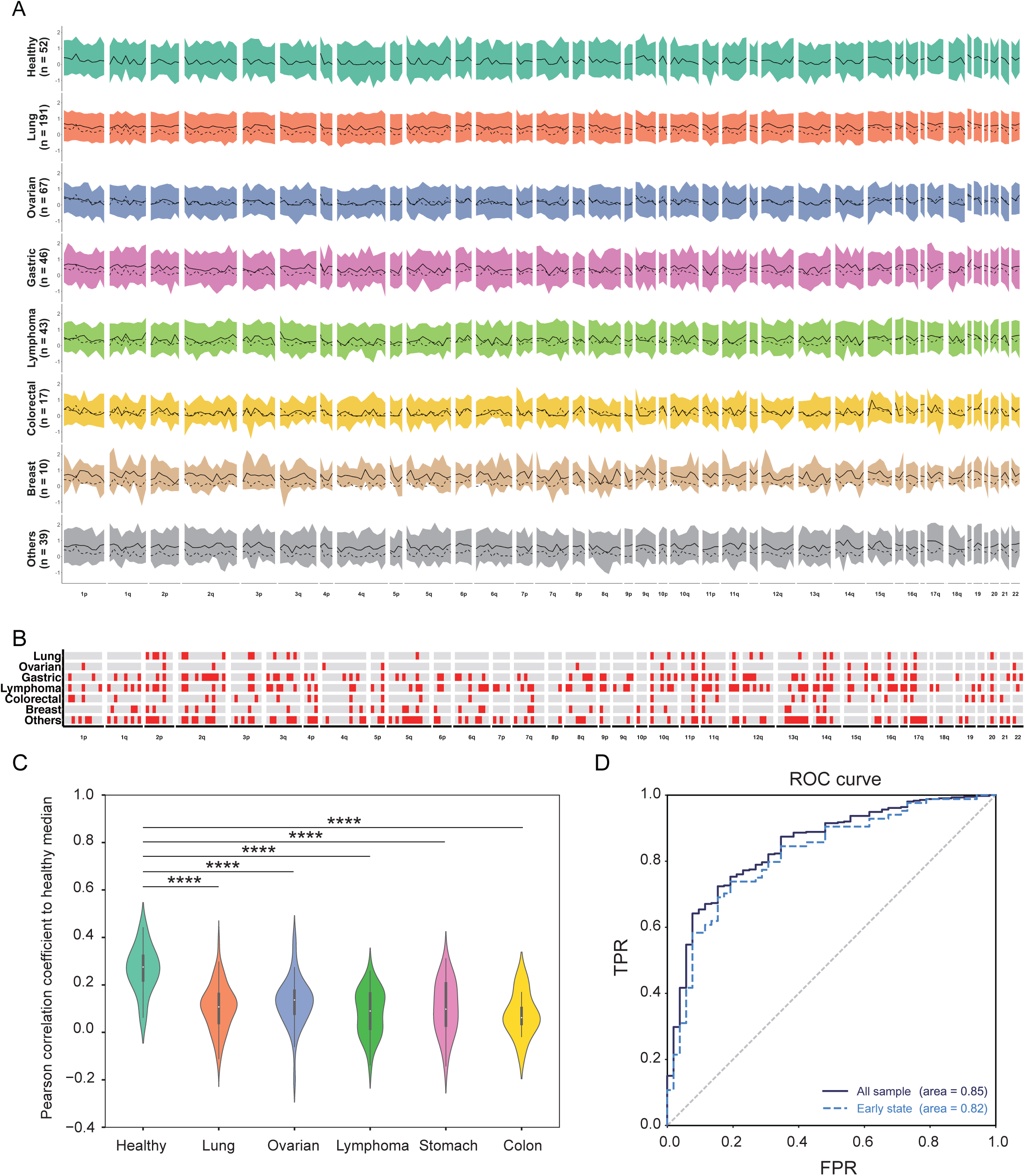
Detection of cancer with ScanTecc. a. short/long cell-free eccDNA profiles from ScanTecc for healthy individuals and patients with 8 cancer types are depicted per cancer type using 10-Mb windows. For each sample, median profile is indicated in black. b. Windows are indicated in red if more than 10% of the cancer samples had a fragment ratio more than three standard deviations from the median healthy fragment ratio. c. The performance of the ScanTecc identifed cancer patients from healthy individuals (AUC =0.85), and early-state cancer patients from healthy individuals (AUC =0.82).This approach is depicted for a cohort of 52 healthy individuals and 413 patients with cancer, including 84 cancer patients at early state (stages I and II)

Next, we used the eccDNA length distribution profiles in Fig. 2B and the fragment profiles in Fig. 3A as inputs for ScanTecc and integrated the XGboost algorism^29^ into ScanTecc, making it a convenient diagnostic model to classify cancer patients from healthy individuals. We assessed the performance of ScanTecc in screening cancer patients by 3-fold cross-validation repeating 30 times and found that ScanTecc successfully detected 270 out of 413 cancer patients (65.4% sensitivity) and misclassified 6 out of 52 healthy individuals (88 % specificity). Receiver operator characteristic analyses for the detection of cancer patients had an area under the curve (AUC) value of 0.85. Notably, ScanTecc detected 49 out of 84 (58.3%) cancer patients at an early stage (stages I and II) with an AUC score of 0.82 (Fig. 3D), suggesting the capability of ScanTecc in screening cancer patients at early stages.

To further explore whether eccDNA can serve as a biomarker in monitoring disease progression after drug treatment, we investigated the alterations in terms of the number of eccDNAs before and after drug treatment in a cohort of 18 patients with lung cancer. After receiving 3∼4 cycles of chemotherapy, immunotherapy, or targeted therapy (Supplementary Table S2), 9 of the 18 patients were identified as partial response (PR) while the other 9 patients were identified as stable disease (SD) or progress disease (PD). We found that the PR patients had a significantly decreased number of eccDNAs than those SD or PD patients (Supplementary Fig. 4B). Taken together, our results suggested that cell-free eccDNAs can be effective biomarkers for tumor onset and progression, and ScanTecc is potentially applicable for the early detection of cancer patients.

### ScanTecc distinguishes primary cancer types by integrating eccDNA enriched gene signatures

To explore the potential capability of ScanTecc in identifying different cancer types, we built a gene-based model based on the genomic charactersitics of the indentified cell-free eccDNA in each sample. We first aligned each indentified eccDNA onto the reference human geneome, and then for each gene defined the number of eccDNA fragments mapped to the gene locus as the gene cyclization probability (GCP) of this gene (see Methods) (Supplementary Fig. 5A-C). We then performed a pair-wise differential analysis of the GCPs between 4 cancer types in which the number of patient samples surveyed was> 20 (particularly, the lung, ovary, gastric cancer, and lymphoma, n = 131, 47, 27, and 26, respectively) and healthy individuals (n=38). We thereby identified a total of 349 significantly differential genes, and unsupervised hierarchical clustering of the GCP matrix grouped these genes into 4 distinct clusters, which clearly distinguishes cancer types from each other as well as from healthy individuals (Fig. 4A). Genes known to be critical drivers for cancer development, including *PFKP*^30^, *BRAF*^31^, *CDH17*^32^, *ASH1L*^33^, were significantly enriched in lung, ovary, gastric cancer, and lymphoma (*P* < 0.05), respectively (Fig. 4B). We also discovered that many known oncogenes were differently but specifically enriched in 4 cancer types (Supplementary Fig. 5D). For example, *NRG1* was enriched in lung cancer and gastric cancer samples, and *WWOX* was enriched in ovary cancer samples, making them as potential signature genes for cancer type identification.

**Fig. 4.**
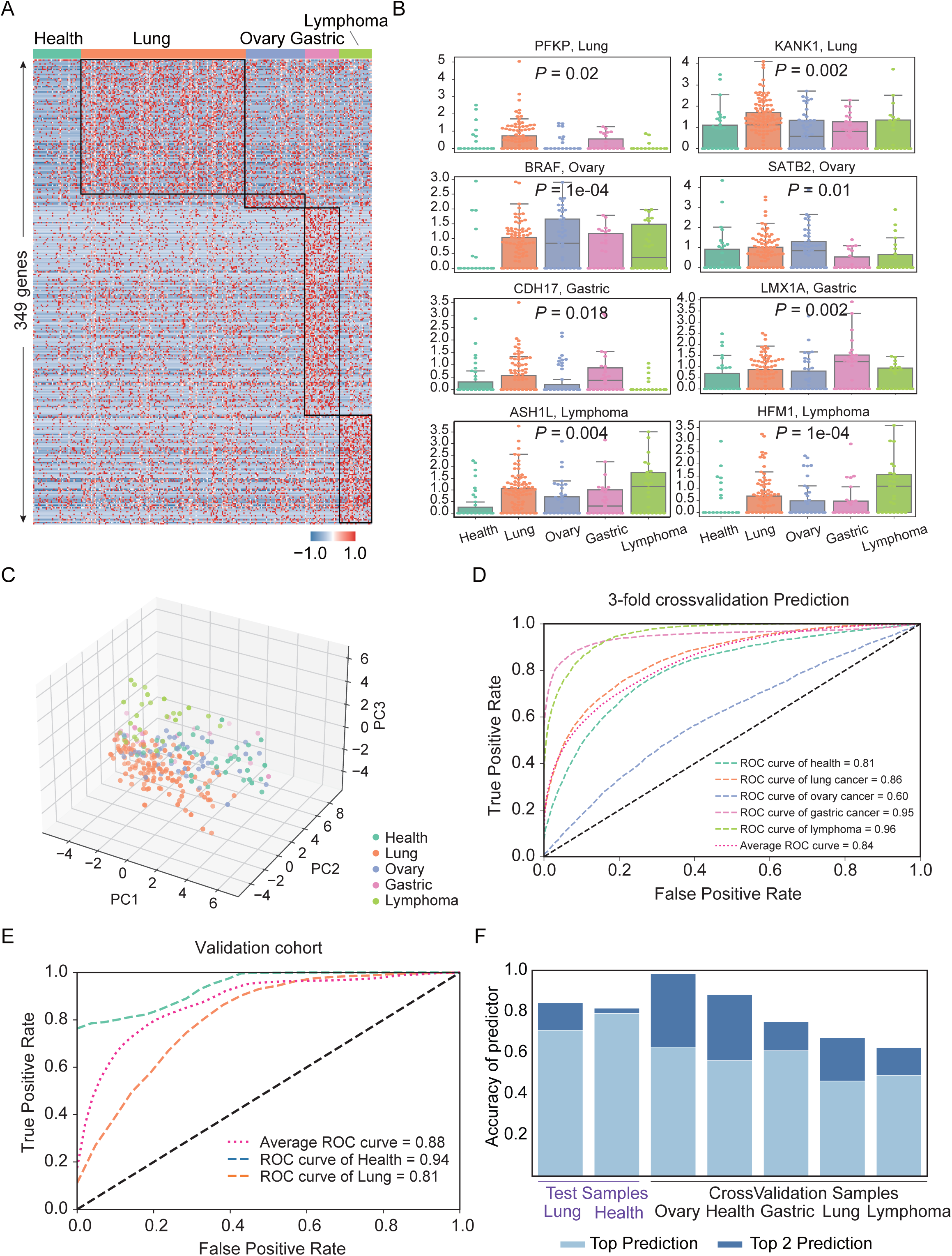
Distinguish of cancer types with ScanTecc. a. Heatmap of 345 significant cell-free eccDNA enriched gene in healthy individuals, lung cancer, ovary cancer, gastric cancer, and lymphoma. b. Boxplot of gene looping probability score of 8 example gene selected from (**a**). c. The first three dimension of principal components analysis (PCA) based on the GLPs of the genes from (a). d. The performance of the ScanTecc distinguishing primary cancer types from healthy individual. e. The performance of the ScanTecc distinguishing lung cancer patients from healthy individual in a independent validation set, including 14 healthy individuals and 21 lung cancer patients. f. Percentages correspond to the proportion of patients correctly classified by one of the two most likely types (sum of light and dark blue bars) or the most likely type(light blue bar) using ScanTecc.

To further examine whether these signature genes can indeed be used as biomarkers to predict cancer types, we performed a principal components analysis (PCA) based on the GCPs of the signature genes and found that different cancer types can be well distinguished (Fig. 4C). We then adopted a widely used machine learning method, support vector machine (SVM)^34^ into ScanTecc to classify all the patients based on the GCPs of these signature genes. We conducted 3-fold cross-validation on ScanTecc predictions and found that the classification of cancer patients had an AUC value of 0.84, ranging from 0.60 to 0.96 (Fig. 4D). To further evaluate the prediction accuracy of our model, we applied ScanTecc to predict lung cancer patients in a subsequent independent validation cohort (14 healthy samples and 21 lung cancer samples) based on the signatures genes from Fig. 4A, and obtained a high prediction accuracy (AUC: 0.88) (Fig. 4E). By integrating this predictive model, ScanTecc can localize the samples into the correct cancer types with accuracy >0.6 (Fig. 4F), and is thereby applicable for the detection of cancers at early-stages (Supplementary Fig. 5E).

## Discussion

EccDNA is commonly characterized in cancer cell lines and somatic tissues. However, there is a lack of studies in a large cohort of cancer patients to characterize the molecular features and functions of eccDNA in peripheral blood. In this study, we demonstrated that cell-free eccDNAs are commonly present in peripheral blood of multiple cancer patients and healthy individuals. These eccDNAs were identified to show greater number and longer fragment size in cancer patients compared with those in healthy individuals. We also discovered that genome-wide fragmentation profiles of eccDNA are different between cancer patients and healthy individuals. Integrated with machine learning methods, we developed ScanTecc and have shown that it can be used to distinguish cancer patients from healthy individuals based on the genomic characteristics of eccDNAs. ScanTecc can also distinguish different tumor types based on the gene cyclization probabilities. Our study provides a non-invasive approach with clinical application potential for the early detection of cancers.

Recent studies have shown that circular DNA is prevalent in human tissues and can be identified by several strategies^4,24^. AA algorithm reconstructs the structure of focally amplified regions using whole-genome sequencing (WGS) data^1^; this approach could detect a large tumor-specific circular DNA catalog that showed a preference of long circle length (>1,000bp). By contrast, Circle-seq and its derivative methods are more sensitive approaches that could obtain a comprehensive characterization of circular DNAs^6^, which is more appropriate for the detection of eccDNA in peripheral blood. Indeed, Kumar et al. reported that microDNAs are present in circulation and detectable in plasma from human^20^. Zhu et al. demonstrated that the length of the majority (≥80%) of cell-free eccDNAs is smaller than 5000bp^21^. In line with these studies, we applied a Circle-seq-based method for eccDNA identification and have shown that this method can detect all the eccDNAs identified by the WGS combined AA strategy, highlighting the high efficiency and sensitivity of the Circle-seq-based method for eccDNA purification and identification in peripheral blood samples.

Oncogene-enriched amplified circular DNA exists in multiple cancer types; these ecDNAs frequently occur in cancer but not in blood or normal tissue. It has been reported that an average of 30%-50% samples from patients with cancer have ecDNAs, which are identified by discordant amplicons and characterized by long fragments and copy number amplification^5,9^. Meanwhile, Circle-seq method identified a large number of short fragment eccDNAs (on average 5,673) per neuroblastoma, and these eccDNAs are not copy number amplified^6^. Moller et al. reported that ∼ 100,000 unique eccDNA types were identified as a result of mutations in muscle and blood samples from 16 healthy men and the majority of these eccDNAs are smaller than 25 kb^26^. These evidences demonstrated that short fragment eccDNA is commonly present in cancer and normal cells.

Previous studies reported the presence of eccDNAs in human plasma from a small number of patients, particularly 23 cancer patients^20^, 3 healthy individuals^21^, and 5 cases of maternal subjects^22^. Here, we have shown that eccDNA profiles in peripheral blood can be used for cancer screening on a large cohort of cancer patients and healthy individuals. However, the biogenesis and functional importance of short fragment eccDNAs still need to be further investigated.

## Data Availability

All data produced in the present study are available upon reasonable request to the authors

## Acknowledgments

This work was supported by the National Key R&D Program of China (2020YFA0112200 to K.Q.), the National Natural Science Foundation of China grants (T2125012, 91940306, 31970858, and 31771428 to K.Q.; 32100457 to J.F.), CAS Project for Young Scientists in Basic Research YSBR-005 (to K.Q.), Anhui Province Science and Technology Key Program (202003a07020021 to K.Q.) and the Fundamental Research Funds for the Central Universities (YD2070002019, WK9110000141, and WK2070000158 to K.Q.). We thank the USTC supercomputing center and the School of Life Science Bioinformatics Center for providing computing resources for this project.

## Author contributions

K. Q., J. F., and C. G. conceived and supervised the project. J. F. designed the framework and performed data analysis with the helps from Q. Y., W. Z., J. Y., K. L., G. X., and X. Y.. S. L., and Y. S. performed atomic-force microscopy and high throughput sequencing experiments with the help from R. S.. Z. Z., J. W., K. D., B. S. provided clincal blood samples from multiple cancers and healthy individuals. C. G., J. F., and K. Q. wrote the manuscript with inputs from all authors. All authors read and approved the final manuscript.

## Competing interests

Jingwen Fang is the chief executive officer of HanGene Biotech. Kun Qu and Chuang Guo are science advisers of HanGene Biotech. The other authors declare that they have no conflict of interest.

## Materials and methods

### Human samples

Peripheral blood samples were collected from cancer patients and healthy control individuals at the First Affiliated Hospital of University of Science and Technology of China, the First Affiliated Hospital of Anhui Medical University, and the Cancer Hospital of The University of Chinese Academy of Sciences. Informed consent was obtained from the patients. Study procedures were followed in accordance with protocols approved by the ethics committee of the University of Science and Technology of China. Detailed clinical information for the patients is described in Supplementary Table 1.

### Sample processing

Peripheral blood samples were collected and centrifuged at 1,600g for 10 min at 4 °C. The plasma portion was further centrifuged at 16,000g for 10 min at 4 °C to remove residual cells and debris. Plasma samples were stored at -80°C. Plasma DNA extractions were performed using QIAamp Circulating Nucleic Acid Kit (Qiagen, 55114).

### EccDNA library preparation and sequencing

For elimination of linear DNA and enrichment of eccDNA, 25 ng of plasma DNA were treated with 5 units of exonuclease V (New England Biolabs) in a 50μL reaction system at 37 °C for 30 min, followed by column purification using MinElute Reaction Cleanup Kit (Qiagen, 28206). Exonuclease digestion and column purification process were performed twice to further eliminate linear DNA. Exonuclease was then heat-inactivated at 70°C for 30 min. Illumina sequencing libraries for eccDNA were prepared by Tn5-transposon-based tagmentation with the TruePrep™ DNA Library Prep Kit V2 (Vazyme, TD501) according to the manufacturer’s instructions. DNA libraries were sequenced with the Illumina NovaSeq6000 platform in 2 × 150-bp paired-end reads.

### Atomic-force microscopy (AFM) imaging

AFM imaging of DNA was performed in dry mode. Briefly, 1/10 volume of 10× imaging buffer (100LmM NiCl_2_ and 100 mM Tris-HCl, pH 8.0) was added to the sample in a final DNA concentration of 0.6–1.0 ng/μl, and 2–5 μl of the mixture was then spread on a freshly cleaved mica (Ted Pella) surface. After 2 min incubation, the specimen was rinsed twice with 200 μl of 2 mM magnesium acetate, drying before and after the rinses with compressed air. Images were acquired by using ScanAsyst in Air probe on a Dimension icon MultiMode V atomic-force microscope in ‘ScanAsyst -Air mode’ and processed with NanoScope Analysis 2.0.

### Data preprocessing

The raw reads were aligned to the human reference assembly hg19 by Burrows-Wheeler Aligner MEM v.0.7.12 with default parameters. The bam file was sorted by samtools v.0.1.19. Samtools flagstat was used to get the statistical comparison results of bam files. PCR and optical duplicates were marked using Sambamba markdup v.0.6.6 with default parameters. The putative eccDNAs were identified by the pipeline that was modified from CIRCexplorer2 (https://circexplorer2.readthedocs.io/en/latest/). The putative eccDNAs with number of reads including breakpoint > 4 and genomic length < 100000000bp were kept for downstream analysis.

### eccDNA fragment size ratio analysis

The normalized count of long eccDNAs for each sample was defined as follows:

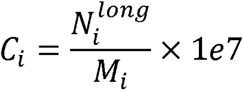

Where 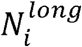 denotes the number of eccDNAs whose genomic length was greater than 10000bp in sample *i*; *M*_*i*_ was the number of mapped reads in sample *i*.

The eccDNAs of each sample was divided into eight bins according to their fragment size: 10^0^∼10^1^bp, 10^1^∼10^2^bp, 10^2^∼10^3^bp, 10^3^∼10^4^bp, 10^4^∼10^5^bp, 10^5^∼10^6^bp, 10^6^∼10^7^bp, 10^7^∼10^8^bp. The length ratio of each bin for each sample was calculated as follows:

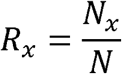

Where *N*_*x*_ denotes the number of eccDNAs whose genomic length is within the bin *x. N* is the total number of eccDNAs.

The length ratio difference between cancer samples and normal samples of each bin was defined as follows:

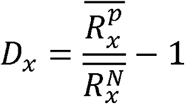

Where 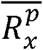 and 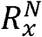 are the average length ratio of the bin *x* in cancer samples and normal samples, respectively. Thus, *D*_*x*_ greater than 0 indicates a greater length ratio of the bin *x* in the cancer samples; *D*_*x*_ less than 0 indicates a greater length ratio of the bin *x* in the normal samples.

### EccDNA genomic distribution analysis

Bedtools was used for the overlap analysis between the eccDNAs and genomic proximal genes. For a given eccDNA, if it does not overlap with any gene, it is defined as an ‘intergenic’ eccDNA; If it falls within any gene, it is defined as ‘full gene’ eccDNA; Otherwise, it is defined as ‘partial gene’ eccDNA. The annotatePeak function of Homer (http://homer.ucsd.edu/homer/) was used for the genomic element analysis of eccDNAs.

### Junction motifs of eccDNA

In order to analyze the motif patterns flanking the eccDNA junctional sites, we scanned the base compositions from 25 bp upstream to 25 bp downstream of the start and end sites for each eccDNA locus. The targeted 50bp sequences around the junctional site of eccDNA molecules were inferred from the reference genome using getfasta function in bedtools. Then we calculated the base frequency of the targeted sequences and visualized the motif patterns by python package logomaker^35^.

### Analyses of autosomal eccDNA fragment size profiles

We tiled the hg19 autosomes into 241 adjacent, non-overlapping 10Mbp windows based on previous work^27^. We excluded regions of low mappability and the Duke blacklisted regions (http://hgdownload.cse.ucsc.edu/goldenpath/hg19/encodeDCC/wgEncodeMapability/). Then, we removed the remainder regions nearby centromeres, whose size is shorter than 10Mbp in each autosome. Using these approaches, we totally removed 471Mbp (16%) of the hg19 reference autosome genome, including centromeric and telomeric regions. The eccDNA fragment size profile of each sample was defined as follows:

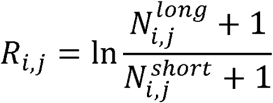

Where *R*_*i,j*_ indicates the fragment ratio of eccDNA in window *j* in the eccDNA fragment size profile of sample *i*. 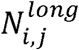 and 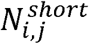 respectively denote the number of eccDNAs in sample *i*, window *j*, which have lengths between 280bp and 500 bp and between 150bp and 280bp.

### Prediction model for sample classification based on eccDNA normalized count

The integrated matrix of eccDNA length distribution profiles and the fragment profiles was used for the XGBoosting prediction model (xgboost v.1.6.1 python package). The prediction model outputs a cancer score for each sample. Cancer scores range from 0 to 1, with higher scores representing higher cancer probability. Each algorithm was cross-validated 3-fold and looped 30 times, and the Receiver Operating Characteristic (ROC) curve was used to plot the model prediction performance and the AUC (Area Under Curve) scores was calculated.

### Cancer type-specific feature extraction

In order to compare the differences of eccDNA fragments across cancer types, we annotated each eccDNA fragment to genes. First, we used the 10kb upstream of the promoter and the gene body region of each gene as gene reference, and the raw count was defined as the number of eccDNA fragments that completely fell within the gene reference. Next, we normalized the raw count by dividing the number of all eccDNA fragments per sample, defined as the gene cyclization probability (GCP). The normalized procedure was defined as:

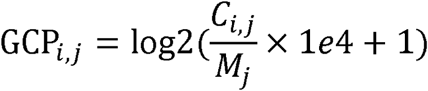

For sample *j, C*_*i,j*_ denotes the number of eccDNA fragments for gene *i*; *M*_*i*_ was the number of eccDNA fragments in sample *j*.

The samples that less than 50% eccDNA fragment completely fell within the gene reference were deleted. Significance analysis was then performed by Wilcoxon Rank Sum Test and genes that were differentially amplified for each cancer type were identified under two conditions: *P* < 0.01, log2 fold change > 0.36. We perform PCA dimensionality reduction on these differential genes, take the top 20 PCs as features and input them into the multi-class model “OneVsRestClassifier” for training. Since the number of lung cancer samples far exceeds other cancer types, we randomly select 30 or 50 lung cancer samples as the training data set, when using 3-fold cross-validation or test sample validation.

### Oncogene enrichment analysis

To investigate which oncogenes play an important role in the formation of eccDNA. We used allOnco (http://www.bushmanlab.org/links/genelists), a set of 2,579 cancer genes generated from curated collections of cancer genes from many different publications. For a given oncogene in a cancer type, if 1) the gene is expressed in less than 50% of healthy samples; 2) the gene is expressed in greater than 50% of cancer samples; 3) the average expression is higher than in healthy samples with Log2Fold > 0.36, the gene was enriched in this cancer type. For a given oncogene in a cancer sample, if the average expression of a gene is higher than in healthy samples with Log2Fold > 0.5, the gene was enriched in this cancer sample.

## FIGURE LEGEND

**Supp Fig. 1.**
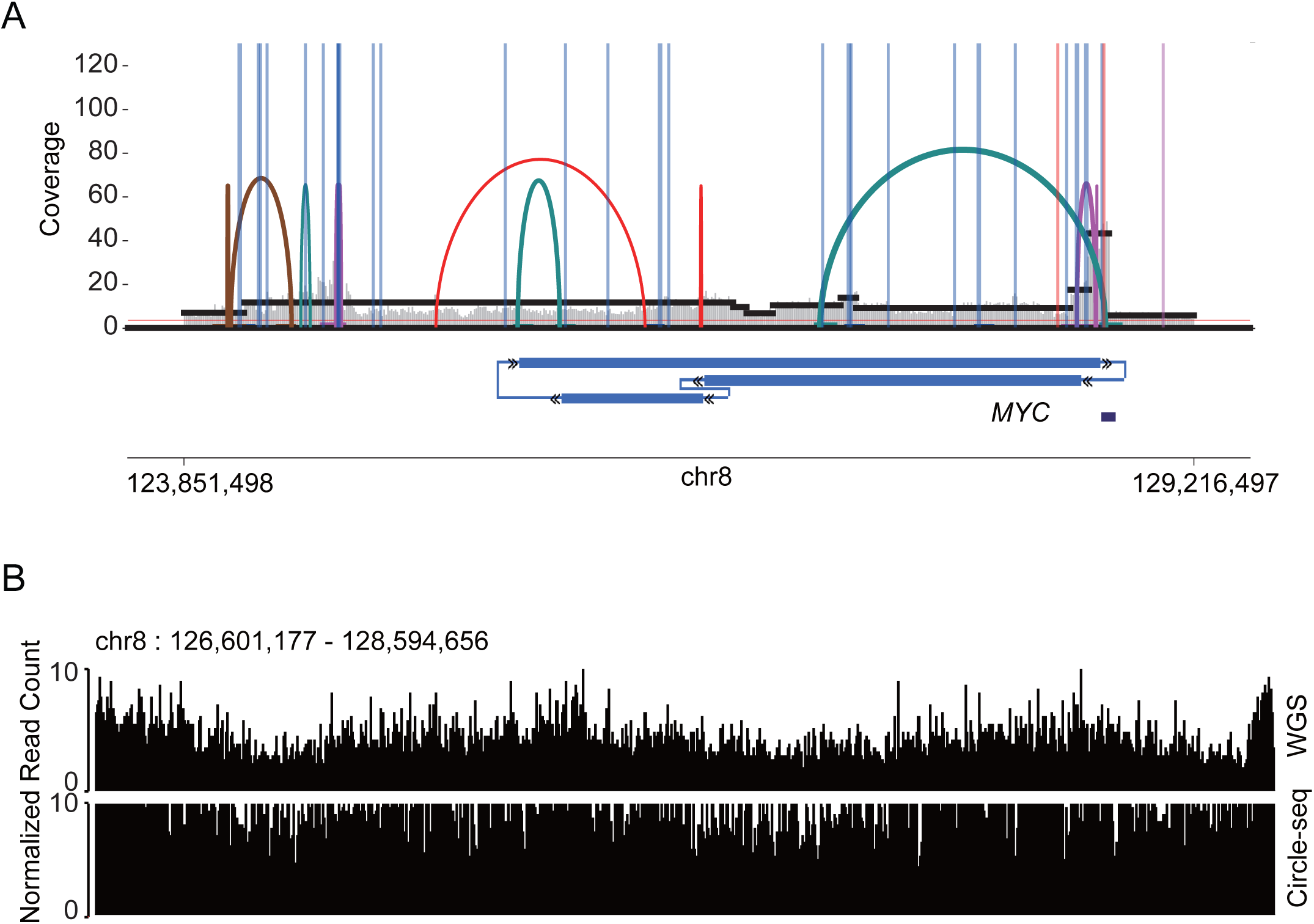
Identification of cell-free eccDNA in blood via WGS & Circle-seq. a. AmpliconArchitect breakpoint graph from bulk WGS of cancer patients blood, showing circular amplicons around MYC. b. Genome coverage of the MYC enriched eccDNA detected via WGS and Circle-seq.

**Supp Fig. 2.**
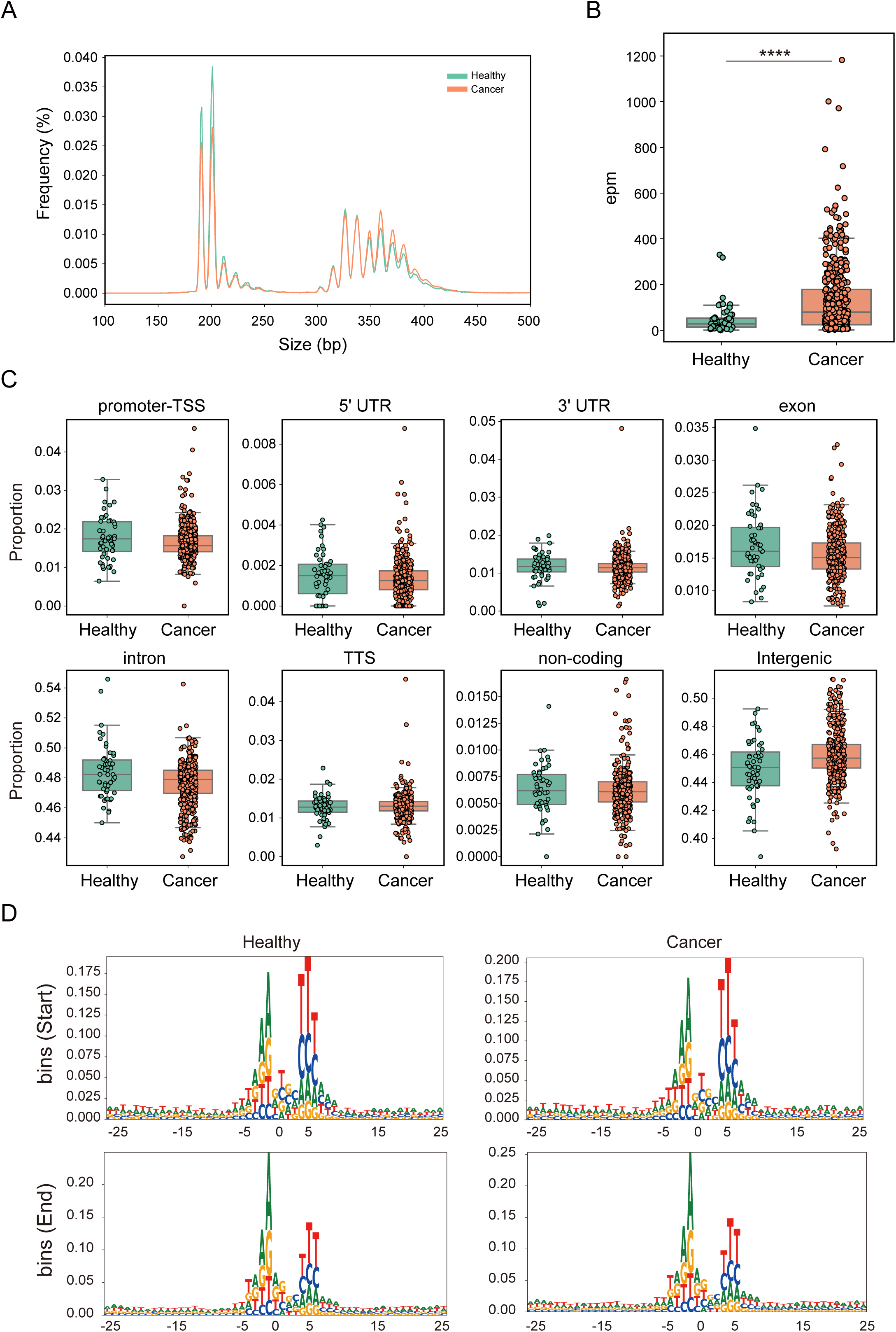
Molecular characteristics of cell-free eccDNA from healthy individuals and cancer patients. a. Size distributions density of cell-free eccDNA from healthy individuals (blue) and cancer patients(orange). b. Normalized cell-free eccDNA counts detected in healthy individuals’ group and all cancer patients. c. Normalized genomic coverage proportion of healthy/patients cell-free eccDNA for each class of genomic elements. d. Trinucleotide motif sequences flanking the start and end positions of healthy/patients cell-free eccDNA molecules with 4-bp spacers in between.

**Supp Fig. 3.**
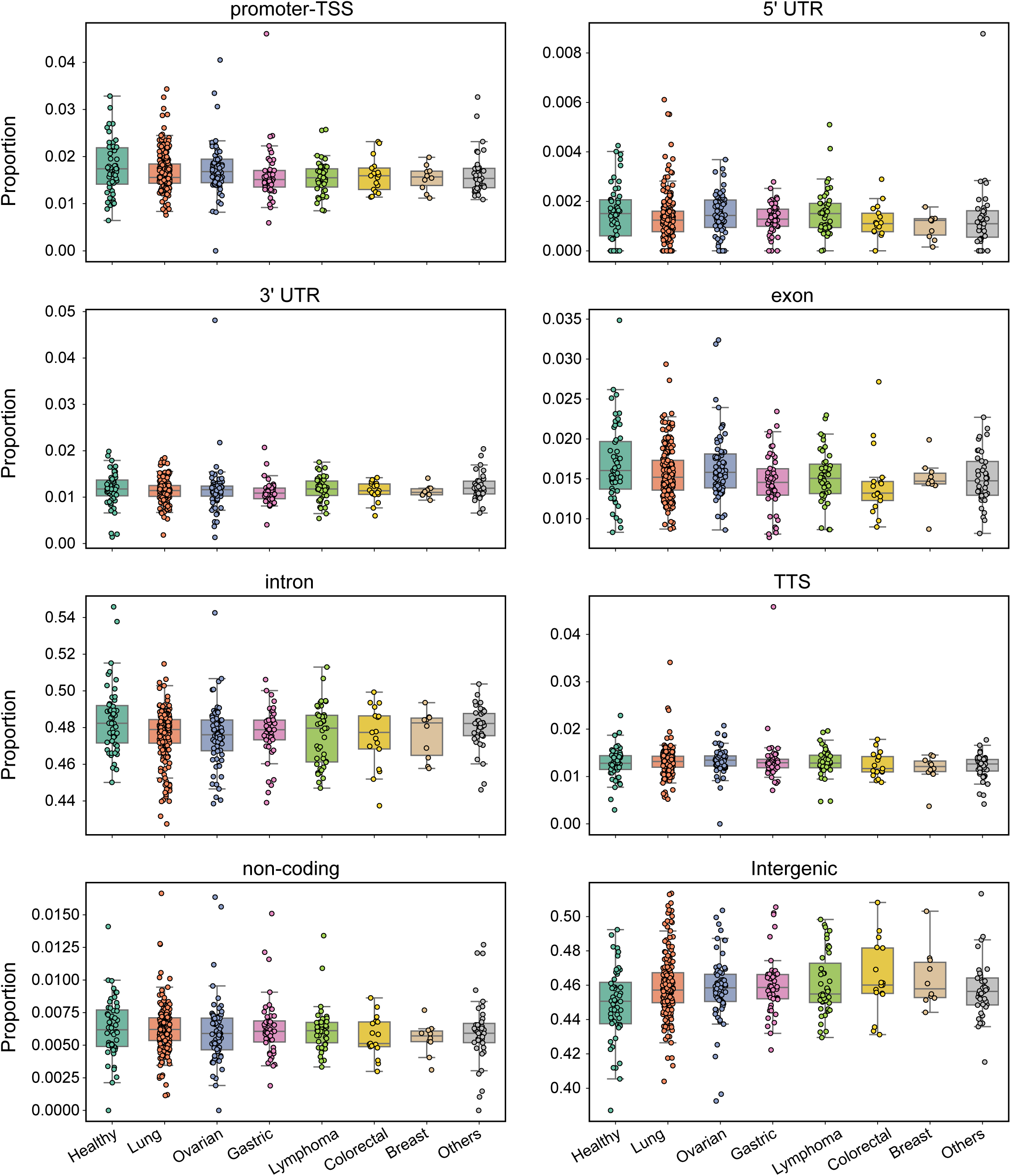
Molecular characteristics of cell-free eccDNA different cancer types patients. a. Normalized genomic coverage proportion of healthy and 8 cancer types patients cell-free eccDNA for each class of genomic elements.

**Supp Fig. 4.**
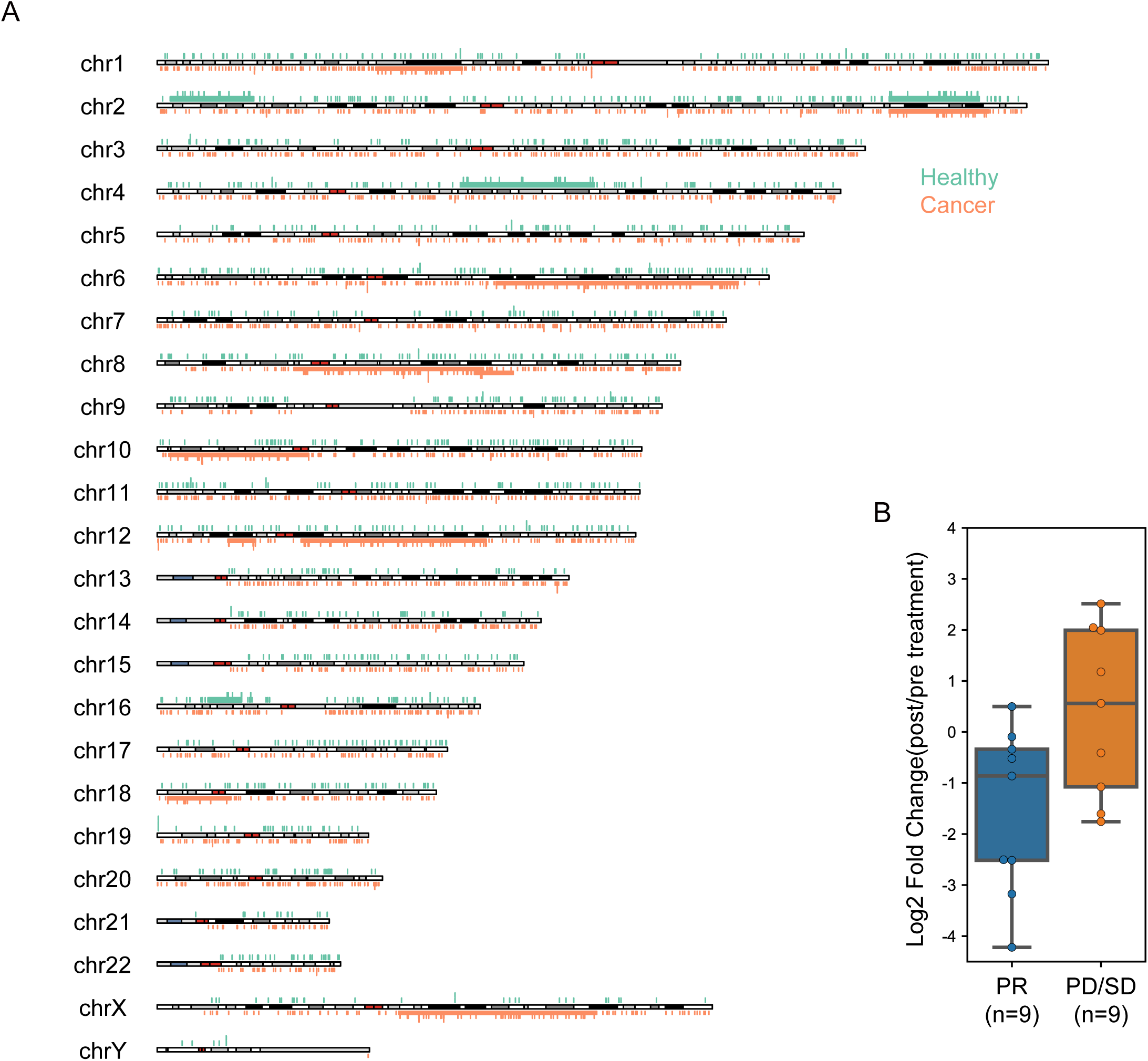
ScanTecc reflects treatment response with change of cell-free eccDNA. a. Karyotype plot showing chromosomal distribution of one healthy individual and one cancer patients. b. Log2 Fold-change of normalized cell-free eccDNA counts per million of pre-therapy/pro-therapy of PR patients(n=9) and SD/PD patients(n=9)

**Supp Fig. 5.**
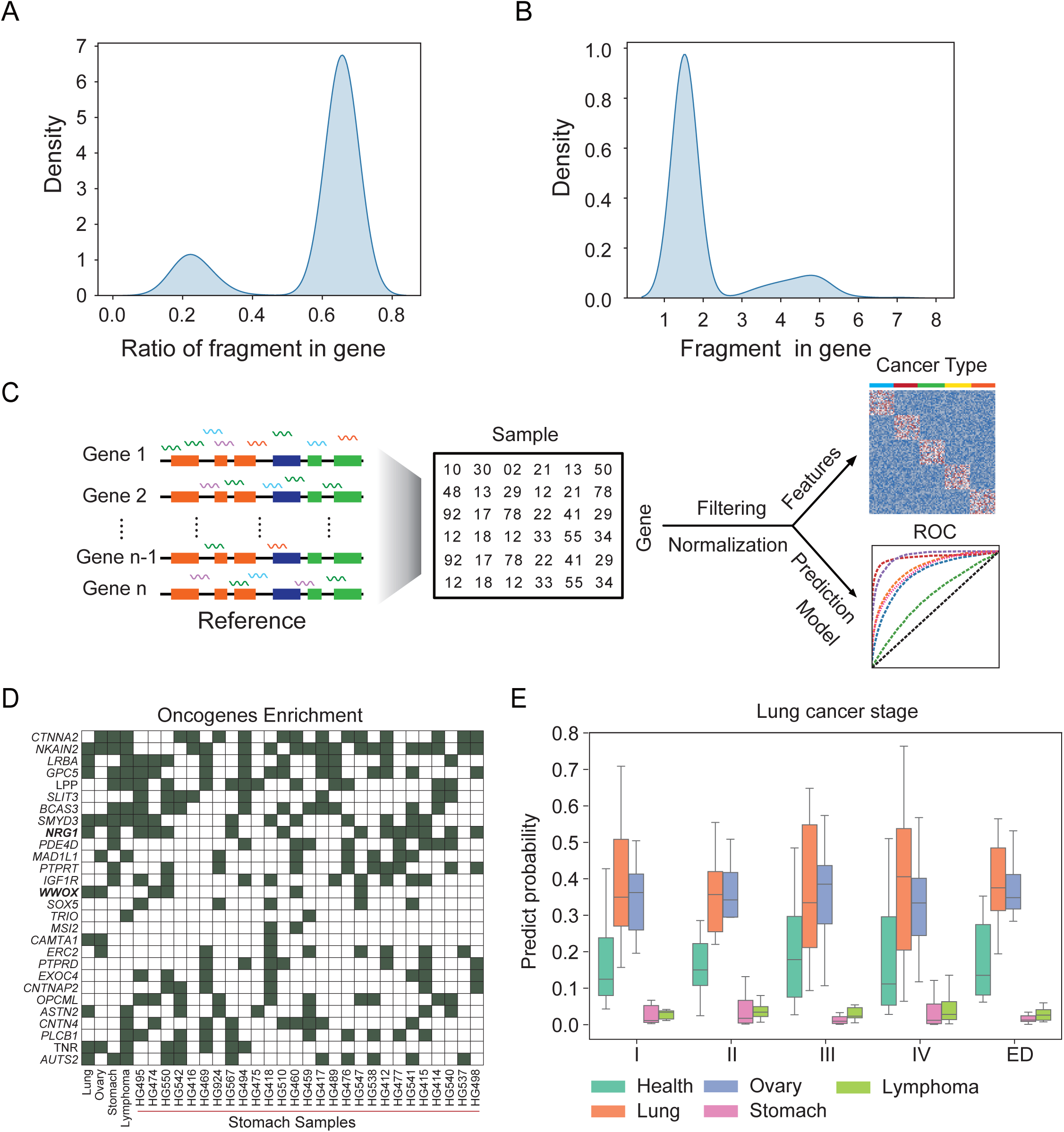
ScanTecc identified cancer type specific enriched gene on cell-free eccDNA through gene looping probability score. a. Percentage of eccDNA fragments in each sample that fall completely within the gene. b. The number of eccDNA fragments in each sample that fall completely within the gene. c. The schema depicts the general workflow of gene looping probability score calculation and machine learning model building. e. The boxplots show the ScanTecc prediction score of lung cancer patients, other cancer type patients, and healthy individuals across different cancer state.

## Notes

### Author Declarations

Ethics committee of the First Affiliated Hospital of University of Science and Technology of China and the First Affiliated Hospital of Anhui Medical University gave ethical approval for this work

