## Supplemental Table 1 for "ScanTecc classifies primary cancers via cell-free extrachromosomal circular DNA in peripheral blood": Table 1_sample_information.docx

**Table 1 Clinical characteristics of cancer and healthy groups**

|  | Cancer (N=413) | Healthy (N=52) | Total (N=465) |
| --- | --- | --- | --- |
| **Age, years** | | | |
| Mean (SD) | 61.37 (12.07) | 50.50 (10.19) | 60.15 (12.35) |
| Median (Q1, Q3) | 63.00 (55.00, 70.00) | 52.50 (42.75, 56.00) | 59.00 (53.00, 69.00) |
| Min, max | 15, 87 | 23, 72 | 15, 87 |
| **Age group, n (%)** | | | |
| <50 years | 54 (13.08) | 18 (34.62) | 72 (15.48) |
| ≥50 years | 359 (86.92) | 34 (65.38) | 393 (84.52) |
| < 65 years | 224 (54.24) | 48 (92.31) | 272 (58.49) |
| ≥65 years | 189 (45.76) | 4 (7.69) | 193 (41.51) |
| **Sex, n (%)** | | | |
| Female | 204 (49.39) | 29 (55.77) | 233 (50.11) |
| Male | 209 (50.61) | 23 (44.23) | 232 (49.89) |
| **Clinical cancer stage, n (%)** | | | |
| I | 46 (11.14) |  | 46 (9.89) |
| II | 38 (9.20) |  | 38 (8.17) |
| III | 105 (25.42) |  | 105 (22.58) |
| IV | 164 (39.71) |  | 164 (35.27) |
| Missing | 60 (14.53) |  | 60 (12.91) |
| **Cancer Type, n (%)** | | | |
| Lung cancer | 191 (46.25) | / | 191 (41.08) |
| Ovarian cancer | 67 (16.22) |  | 67 (14.41) |
| Lymphoma | 43 (10.41) |  | 43 (9.25) |
| Gastric cancer | 46 (11.14) |  | 46 (9.89) |
| Colorectal cancer | 17 (4.12) |  | 17 (3.66) |
| Breast cancer | 10 (2.42) |  | 10 (2.15) |
| Pancreatic cancer | 6 (1.45) |  | 6 (1.29) |
| Adenocarcinoma of the esophago-gastric junction | 3 (0.73) |  | 3 (0.65) |
| Endometrial cancer | 5 (1.21) |  | 5 (1.08) |
| Esophageal carcinoma | 2 (0.48) |  | 2 (0.43) |
| Carcinoma of the fallopian tube | 2 (0.48) |  | 2 (0.43) |
| Soft tissue sarcoma | 2 (0.48) |  | 2 (0.43) |
| Cervical cancer | 4 (0.97) |  | 4 (0.86) |
| Renal cancer | 2 (0.48) |  | 2 (0.43) |
| Hypopharyngeal cancer | 1 (0.24) |  | 1 (0.22) |
| Oral cancer | 1 (0.24) |  | 1 (0.22) |
| Peripheral nerve sheath tumor | 1 (0.24) |  | 1 (0.22) |
| Glioma | 4 (0.97) |  | 4 (0.86) |
| Melanoma | 1 (0.24) |  | 1 (0.22) |
| Liver cancer | 2 (0.48) |  | 2 (0.43) |
| Nasopharyngeal carcinoma | 1 (0.24) |  | 1 (0.22) |
| Cancer of unknown primary site | 1 (0.24) |  | 1 (0.22) |
| Gynecological tumor | 1 (0.24) |  | 1 (0.22) |
